# Estimating Burden of Disease among blind individuals with Non-24-Hour Sleep-Wake Disorder

**DOI:** 10.1101/2020.04.11.20061887

**Authors:** Lauren Van Draanen, Derek Xiao, Mihael H. Polymeropoulos

## Abstract

**Purpose:** To quantify the burden of disease in blind patients with Non-24-Hour Sleep-Wake Disorder (N24HSWD), utilizing longitudinal sleep diary data. N24HSWD is a circadian disorder characterized by a cyclical pattern of aberrant circadian and sleep-wake cycles that are associated with increased frequency of sleep episodes during the school/work day hours. Daytime sleep episodes will decrease the opportunity of school/work participation significantly impacting the quality of life of the patient.

**Methods:** We used the day time sleep diary data from a period of approximately 90 days in blind individuals with a sleep complaint, which were identified from a group of blind individuals with N24HSWD (n=121) and a control group of blind individuals without N24HSWD (n=57).

**Results:** N24HSWD patients had more frequent and longer episodes of daytime sleep as compared to a control group. N24HSWD patients also had significantly fewer healthy days, defined by daytime sleep free days (DSFD), as compared to the control group.

**Conclusion:** Daytime sleep free day (DSFD) is a useful and specific measure of disease burden in patients with N24HSWD and it is predicted to be correlated with the standardized HRQOL-4, Healthy Days measurement.

## Introduction

Non-24-Hour Sleep-Wake Disorder (N24HSWD) is a rare circadian disorder that affects both sighted and blind patients but is highly prevalent among individuals who are totally blind and lack light perception. N24HSWD results from a misalignment of the body’s internal circadian timing system with the external 24-hour clock. In the human brain, a complex circadian timing system residing in the suprachiasmatic nucleus (SCN) sets in motion the coordinated expression of a myriad of physiological processes, including the sleep-wake cycle, hormonal secretion, feeding behavior, and metabolic processes, among many. The endogenous timing system has a beat cycle of a little longer than 24 hours and in blind individuals, it is around 24.25 hours. This endogenous circadian system is reset every day through the perception of light through the retina, therefore maintaining a periodicity of 24 hours. In totally blind individuals with no light perception, this ability to reset the internal circadian system is missing. This results in a constant and progressive misalignment between the internal and external clock. The result is a sleep wake cycle that is misaligned with social norms and in turn, results in a N24HSWD patient sleeping and waking at the wrong times of the day for extended periods, synchronizing for a brief period of time and then the cycle repeats in a perpetual manner. Patients with N24HSWD suffer chronically from the condition, which has significant impact in social and occupational functioning. For the extended periods when the endogenous cycle creates daytime sleep propensity, patients cannot resist falling asleep in the middle of the day, a situation often incompatible with successful performance in school and work activities. In this study we aim to quantify the burden of disease in N24HSWD patients, by utilizing longitudinal sleep diary data and transforming them into a surrogate measurement of healthy days in a given 30-day period. We refer to this measurement as daytime sleep free days (DSFD), measured through different thresholds of sleep between 9:00 AM and 5:00 PM. DSFD is expressed as daytime sleep free days in a 30-day period. This transformation allows us to compare this measure with unhealthy days as measured by the Centers for Disease Control (CDC) Behavriorial Risk Factor Surveillance System Health Related Quality of Life (HRQOL-4) questionnaire in the general population. While further studies will be needed to validate the DSFD as a measure of disease burden in N24HSWD, our work establishes DSFD as a specific tool to allow for the study of the impact of N24HSWD in patients with this severe and debilitating chronic condition.

## Materials and Methods

### Cohort Description

The patients in this study are a sequential cohort of blind patients with a sleep complaint that participated in a clinical program, aimed at the development of a treatment for N24HSWD. The study, known as the Safety and Efficacy of Tasimelteon (SET) study was conducted between 2010 and 2013 in sites in the United States, France and Germany^1^. Participating individuals agreed to the procedures of the study by signing an informed consent, which was approved by the appropriate Institutional Review Boards (IRB). In total for this study we are including 178 blind patients with a sleep complaint, of which 121 were shown to have N24HSWD and 57 did not and are used as controls. Demographic data for the participants are shown in Table 1.

**Table 1.** Characteristics of the control and Non-24-Hour Sleep-Wake Disorder subjects from the SET^1^ study

| Characteristic | Disease Status |  |
| --- | --- | --- |
|  | Control<br>(N=57) | N24HSWD<br>(N=121) |
| Sex |  |  |
| Male | 20 (35%) | 72 (60%) |
| Female | 37 (65%) | 49 (40%) |
| Age (years) |  |  |
| Mean (SD) | 46.9 (13.45) | 50.8 (13.26) |
| Race |  |  |
| American Indian or Alaska Native | 2 (3.5%) | 1 (1%) |
| Asian | 1 (1.8%) | 1 (1%) |
| Black or African American | 9 (16%) | 18 (15%) |
| Native Hawaiian or other Pacific Islander | 1 (1.8%) | 0 (0%) |
| Other | 0 (0%) | 3 (2%) |
| White | 44 (77%) | 98 (81%) |
| Weight (kg) |  |  |
| Mean (SD) | 46.86 (13.45) | 50.82 (13.27) |
| BMI (kg/m<sup>2</sup>) |  |  |
| Mean (SD) | 27.8 (3.79) | 28.15 (3.9) |
| Tau (hours) |  |  |
| Mean (SD) | 23.9 (0.04) | 24.54 (0.27) |
| Average length of sleep diary collection (days) |  |  |
| Mean (SD) | 64.01 (39.2) | 92.82 (49.71) |

The safety and efficacy of tasimelteon to treat Non-24 in totally blind individuals was assessed in two phase 3, randomised, placebo-controlled, double-masked, multicentre trials, SET and RESET (Randomised withdrawal study of the Safety and Efficacy of Tasimelteon). These trials were completed in 27 US and six German clinical research centres and sleep centres. The studies were approved by central and local Institutional Review Boards (the Ethics Committees that reviewed SET protocols in Germany were Ethikkommission der Ärztekammer Hamburg and Landesamt für Gesundheit und Soziales Berlin; the Ethics Committees that reviewed the SET or RESET protocols, or both, in the USA were Chesapeake Research Review, the Institutional Review Board New York Eye and Ear Infirmary, Minneapolis Medical Research Foundation Human Subjects Research Committee, Partners Human Research Committee, the Research Compliance Office Stanford University, and St Luke’s Hospital Institutional Review Board). These studies were done in accordance with Good Clinical Practice, as required by the US Food and Drug Administration and German regulatory bodies (where applicable), and in agreement with the Declaration of Helsinki.

### Sleep Diaries/CDC HRQOL-4 Questionnaire

Daily sleep diary data were collected through an interactive voice response system (IVRS)^1^. Average length of sleep diary collection for the two groups is shown in the demographics table (Table 1). The HRQOL-4 Questionnaire consists of four standardized questions developed by the CDC in 1993 ^2, 3^. The four questions are used to evaluate overall health, physical and mental health. After the first overall health question, the following three questions are used to evaluate how physical health, mental health and both physical and mental health were “not good” in the last 30 days (Table 2). These questions have been validated across the United States and various patient populations as a robust method to measure a person’s quality of life and determine how many days in a month they feel that their physical health, mental health or both affected their days ^4, 5, 6^. In the CDC data from 2013, 2014 and 2015, 1,397,893 individuals participated in the survey.

**Table 2.**
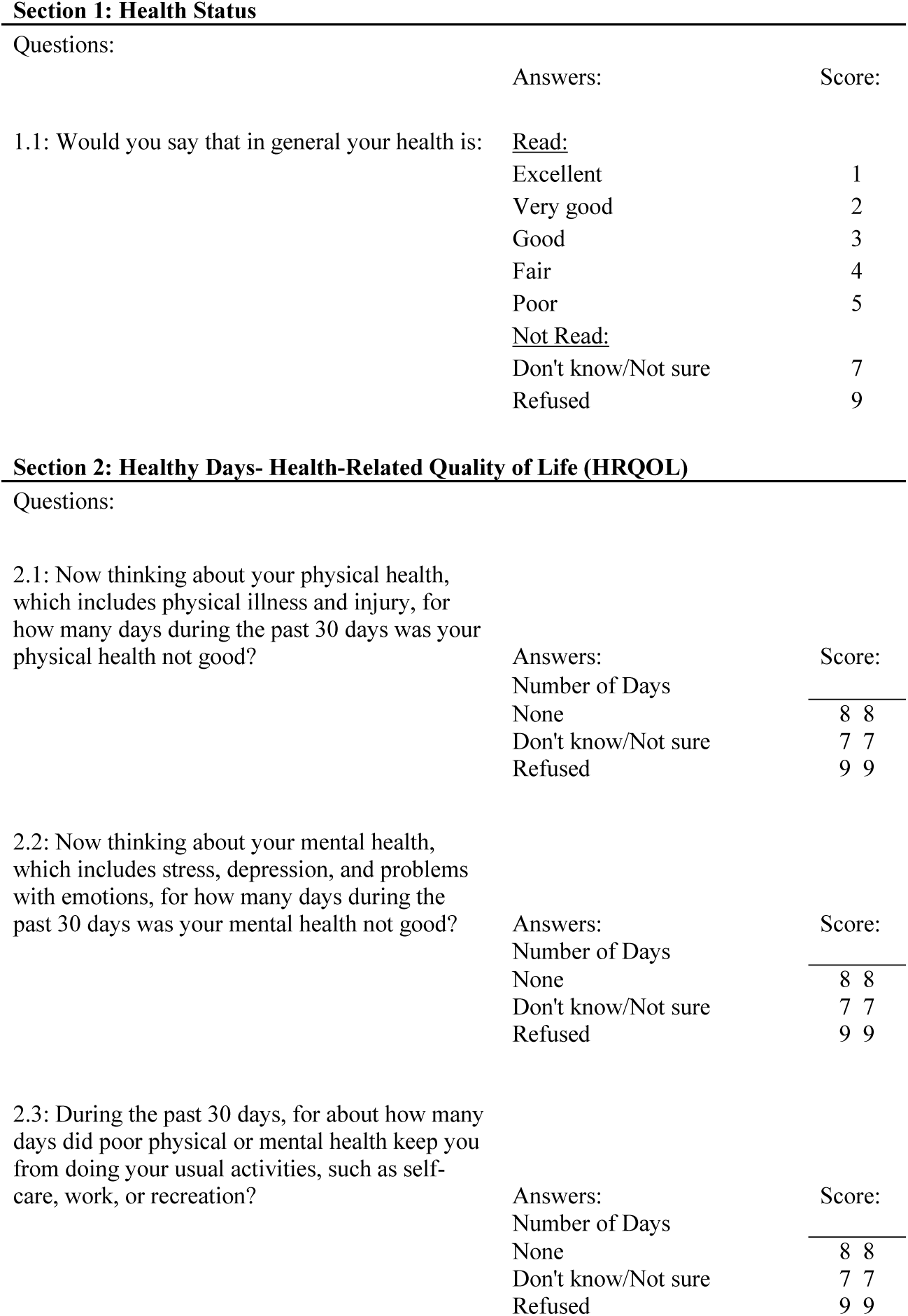
CDC Behavioral Risk Factor Surveillance System Questionnaire (HRQOL)^3^

The HRQOL instrument is used to measure the subjective quality of life for people across diverse geographies ^8-15^. The CDC standardized a HRQOL questionnaire in the early 1990s and it has been validated across the United States and the globe ^8, 10^. The basic questionnaire includes four questions related to mental and physical health, and it has been expanded across various disease states and physical and mental conditions (Table 1). Adaptations have been created in many languages, by large organizations such as the World Health Organization and various countries and states across the world.

### Data Transformations

To assess the disease burden of N24HSWD on the daily social/working life, the daily sleep diary data reported from 9:00 AM to 5:00 PM in an interactive voice recording system (IVRS) were studied. Given the cyclical nature of the disorder and in order to increase the sensitivity of detection of a DSFD difference between N24HSWD patients and controls, the worst quartile of the sleep diary collection was used as previously described and referred to as Upper Quartile Daytime Sleep Duration (UQ-dTSD) (Table 3). UQ-dTSD data was assessed to determine the probability of DSFD per day under different thresholds. The probability of DSFD per day will be transformed to the actual number of DSFD within 30 days (Table 4). DSFDs were calculated using four different threshold criteria. A day without any sleep, a day with less than half an hour of sleep, a day with less than one hour of sleep, or a day with less than two hours of sleep (Table 3). These thresholds were used to determine the number of DSFDs out of a 30-day month. The remaining (non-DSFD) number of days within the 30-day period were compared to the unhealthy days per 30 days from HRQOL-4 (Table 5).

**Table 3.** Summary of the daytime sleep duration (9:00 AM - 5:00 PM) by disease status.

|  | Disease Status |  | *p*Value<sup>3</sup> |
| --- | --- | --- | --- |
|  | Control<br>(N=57) | N24HSWD<br>(N=121) |  |
| Daytime sleep duration (dTSD) |  |  |  |
| dTSD<sup>1</sup> (hours) (SD) | 0.38 (0.45) | 0.60 (0.57) | 0.0006 |
| UQ_dTSD<sup>2</sup> (hours) (SD) | 1.04 (0.95) | 1.85 (1.49) | <0.0001 |

<sup>1</sup> dTSD: average daytime sleep duration (hours) between 9:00 AM and 5:00 PM.
<sup>2</sup> UQ\_dTSD: upper quartile daytime sleep duration (hours).
<sup>3</sup> p-values are based on an ANCOVA model.

**Table 4.** Summary of the daytime sleep free days (DSFD)^1^.

|  | Disease Status |  | *p*Value<sup>3</sup> |
| --- | --- | --- | --- |
|  | Control<br>(N=57) | N24HSWD<br>(N=121) |  |
| Threshold for daytime sleep free days (DSFD) |  |  |  |
| 0 hours of daytime sleep |  |  |  |
| DSFD/Total<sup>2</sup>: mean (SD) | 0.31 (0.37) | 0.18 (0.28) | 0.0050 |
| DSFD/month (days) | 9 | 5 |  |
| 0.5 hour of daytime sleep |  |  |  |
| DSFD/Total: mean (SD) | 0.42 (0.37) | 0.24 (0.31) | 0.0006 |
| DSFD/month (days) | 13 | 7 |  |
| 1 hour of daytime sleep |  |  |  |
| DSFD/Total: mean (SD) | 0.58 (0.38) | 0.37 (0.33) | <0.0001 |
| DSFD/month (days) | 17 | 11 |  |
| 2 hours of daytime sleep |  |  |  |
| DSFD/Total: mean (SD) | 0.78 (0.35) | 0.60 (0.36) | <0.0001 |
| DSFD/month (days) | 23 | 18 |  |

<sup>1</sup> DSFD are the number of daytime sleep free days in a 30-day period. Four different thresholds define daytime sleep free days: 0, 0.5, 1, and 2 hours of daytime sleep between 9:00 AM and 5:00 PM.
<sup>2</sup> DSFD/Total is the number of daytime sleep free days that are at or below the threshold divided by the total number of diary recorded days.
<sup>3</sup> p-values are based on an ANCOVA model.

**Table 5.**
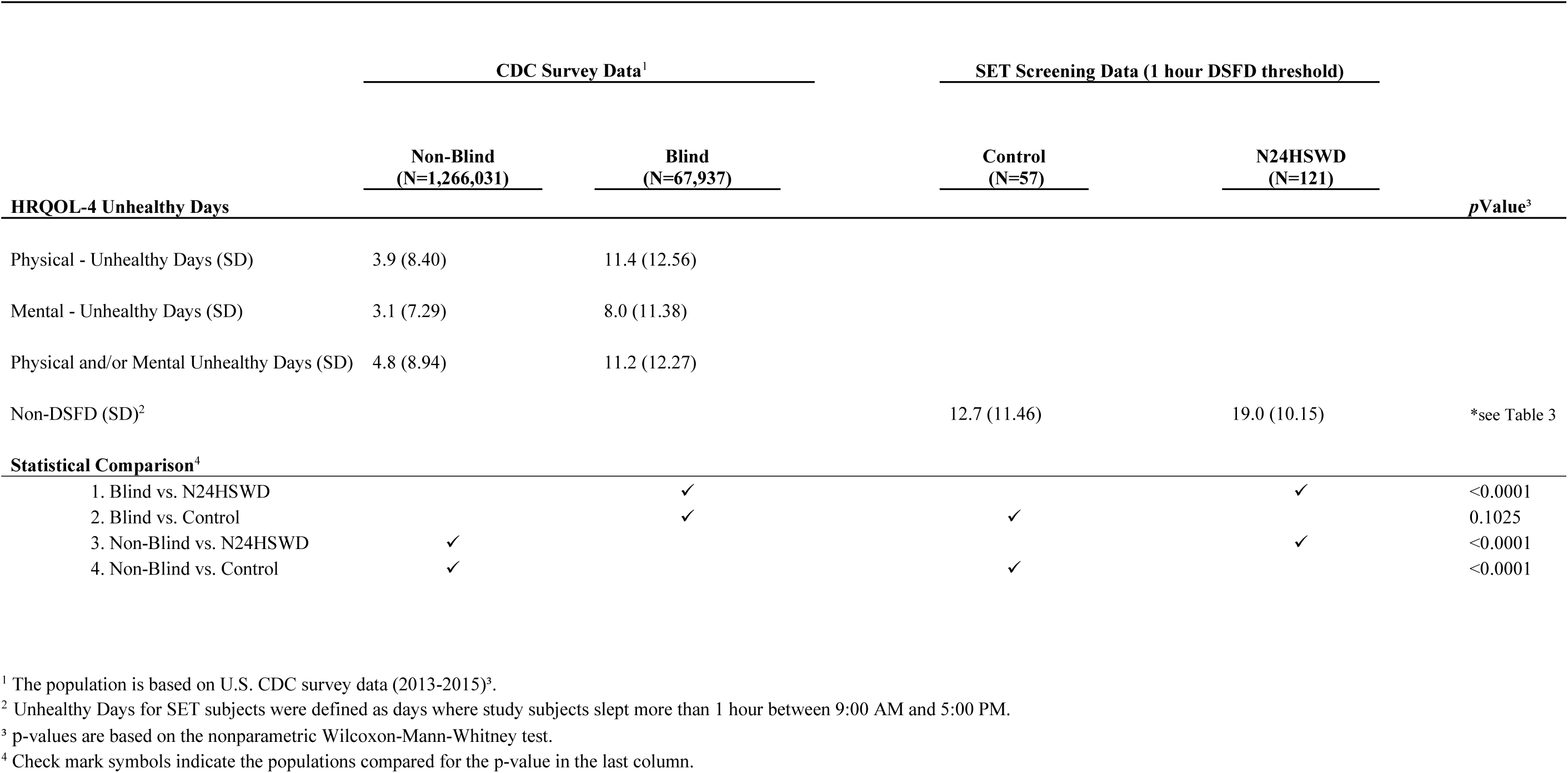
Comparison of daytime sleep free days in the SET study and CDC Health Related Quality of Life (HRQOL-4) survey responses in the U.S. population (blind and non-blind).

|  | CDC Survey Data<sup>1</sup> |  | SET Screening Data (1 hour DSFD threshold) |  |  |
| --- | --- | --- | --- | --- | --- |
|  | Non-Blind<br>(N=1,266,031) | Blind<br>(N=67,937) | Control<br>(N=57) | N24HSWD<br>(N=121) |  |
| HRQOL-4 Unhealthy Days |  |  |  |  | *p*Value<sup>3</sup> |
| Physical - Unhealthy Days (SD) | 3.9 (8.40) | 11.4 (12.56) |  |  |  |
| Mental - Unhealthy Days (SD) | 3.1 (7.29) | 8.0 (11.38) |  |  |  |
| Physical and/or Mental Unhealthy Days (SD) | 4.8 (8.94) | 11.2 (12.27) |  |  |  |
| Non-DSFD (SD)<sup>2</sup> |  |  | 12.7 (11.46) | 19.0 (10.15) | *see Table 3 |
| Statistical Comparison<sup>4</sup> |  |  |  |  |  |
| 1. Blind vs. N24HSWD |  | ✓ |  | ✓ | <0.0001 |
| 2. Blind vs. Control |  | ✓ | ✓ |  | 0.1025 |
| 3. Non-Blind vs. N24HSWD | ✓ |  |  | ✓ | <0.0001 |
| 4. Non-Blind vs. Control | ✓ |  | ✓ |  | <0.0001 |

<sup>1</sup> The population is based on U.S. CDC survey data (2013-2015)<sup>3</sup>. <sup>2</sup> Unhealthy Days for SET subjects were defined as days where study subjects slept more than 1 hour between 9:00 AM and 5:00 PM. <sup>3</sup> p-values are based on the nonparametric Wilcoxon-Mann-Whitney test. <sup>4</sup> Check mark symbols indicate the populations compared for the p-value in the last column.

### Statistical Analysis

An ANCOVA model adjusted for demographic variables (sex, age, race and BMI) was used to assess the difference of daytime sleep duration (dTSD), UQ-dTSD, and DSFD between N24HSWD patients and controls. When comparing to the U.S. CDC HRQOL-4 data, a nonparametric test of Wilcoxon-Mann-Whitney was used. The p-values were displayed to assess the significance at 0.05 levels.

All analyses were conducted using SAS 9.4 (SAS Institute Inc., Cary, NC, USA).

## Results

The N24HSWD patients showed a statistically significant higher dTSD and UQ-dTSD within the normal working/school time period than the control cohort (Table 3). We defined “daytime” as the time between 9:00 AM and 5:00 PM, as these are typical hours used for most work or school days and therefore extended sleep duration during that time would likely interfere with standard work or school activities. dTSD and the UQ_dTSD for control patients was 0.38 or 1.04 hours, while it was significantly higher at 0.60 or 1.85 hours for N24HSWD patients (p-values are 0.0006 and <0.0001, respectively). Blind patients with N24HSWD had more sleep burden during the working /school time than the control cohort of blind patents without N24HSWD. Prolonged periods of daytime sleep are often seen among patients suffering with N24SWD. This is due to misalignment of their circadian rhythm with the 24-hour day ^1, 7^. These episodes of daytime sleep are highly disruptive in the social and occupational functioning of these patients. In most work or school environments sleep episodes during the typical 9:00 AM to 5:00 PM schedule are incompatible with successful employment or school performance. While any length of a sleep episode is disruptive, for the purposes of the work presented in this manuscript we developed four categorical thresholds that defined Daytime Sleep Free Days (DSFD) as days with no daytime sleep, less than a half an hour of sleep, less than one hour of sleep or less than two hours of sleep. N24HSWD patients had consistently lower DSFD than the control cohort of blind patents without N24HSWD for all thresholds (Table 4, all p < 0.01). If we considered a day with 1 hour or more of daytime sleep during the working/social time as an ‘unhealthy day’ within the study population, Table 5 summarizes the comparisons between the study populations and the U.S. CDC survey populations (2013-2015) regarding the HRQOL-4 survey of ‘unhealthy days’. Blind patients with N24HSWD had a significant higher number of unhealthy days than the blind population in the CDC data (p <0.0001). In contrast to the N24HSWD cohort, blind patients without N24HSWD (control cohort) showed a similar number of unhealthy days as compared to the blind population in the CDC data (12.7 versus 11.2 respectively, p=0.1025). Finally, blind patients with N24HSWD and without N24HSWD (control cohort) had much higher numbers of unhealthy days than the non-blind population in CDC (both p<0.0001).

## Discussion

People with self-reported blindness in the U.S. CDC survey population had approximately the same number of unhealthy days as the control patients from the SET study. N24HSWD patients had significantly more unhealthy days than both the control population and the blind and non-blind populations of the U.S. CDC survey populations. This demonstrates the additional burden that blind patients with N24HSWD are experiencing, specifically due to the sleep-wake cycle misalignment induced by the condition.

N24HSWD results in significant impairment, in part due to daytime sleep propensity that results in a number of days within a given month where daytime sleep occurs during the 9:00 AM to 5:00 PM timeframe. The burden of sleep during the day prevents individuals with capacity from effectively functioning within their communities. In addition to the actual daytime sleep, we can assume that N24SWD patients would also experience significant tiredness that can be debilitating as well. Our study shows that the daytime impairment is more significant among N24HSWD patients than in patients of the control cohort, which share similar baseline characteristics and who present with a sleep complaint.

Under the assumption that one hour of daytime sleep significantly affects function, we defined DSFD as days with less than one hour of daytime sleep. Under this condition N24HSWD patients experienced 11 DSFD, compared to patients without N24HSWD who presented with 17 DSFD (p<0.0001). The number of non-DSFD became the unhealthy days, indicating one hour or more of daytime sleep. This exhibits the additional difficulties that patients with N24HSWD face. The N24HSWD patients had significantly more unhealthy days than the control cohort (19 versus 13 respectively, p<0.0001).

In order to examine how the magnitude of DSFD compared with impairment in the population, we used the annual U.S. CDC data collected through the Health Related Quality of Life survey (HRQOL). We made the assumption that Healthy Days as defined in the HRQOL are equivalent to DSFDs, for the purposes of analysis. The HRQOL is utilized to measure a person’s subjective quality of life. We used CDC data from 2013-2015 to analyse “Healthy/Unhealthy Days” in the general population, as well as the self-reported visually impaired subpopulation. N24HSWD patients experienced fewer “healthy days” in a 30-day period than the general U.S. population in the CDC survey (Table 5). This is to be expected, with N24HSWD patients facing the burden of total blindness as well as a non-entrained circadian rhythm. Additionally N24HSWD patients experienced fewer “healthy days” in a 30-day period than patients with self-reported visual impairment in the CDC survey. In addition to the significant difference between the DSFD for N24HSWD patients and control cohort patients from the SET study, there is a significant difference between the “healthy days” for N24HSWD patients and the visually impaired U.S. CDC population. This again exhibits the additional burden of disease that N24HSWD presents to patients. The “healthy days” that control patients and CDC visually impaired patients experienced were not statistically significant, as we might expect. Control cohort patients were found to have an entrained circadian rhythm, despite sleep complaints, which may be a reason that they appear more similar to the CDC visually impaired patients, and they are different from N24HSWD patients. We conclude that DSFD can be a useful measure of impairment in patients with N24HSWD disorder and could allow for the evaluation of the impact of treatments aimed at improving the symptoms of this disorder.

There are several limitations of this work that may impact the conclusions. First, N24HSWD patients did not receive the HRQOL questionnaire and therefore direct comparison cannot be made. Second, the CDC population described visual impairment, likely includes patients who are not totally blind, in contrast to the totally blind population of the SET study. Notwithstanding these limitations it is likely that directionally our conclusions are valid, while the magnitude of the comparative observations will be less certain.

Future studies could be designed to prospectively quantify the impact of N24SWD on quality of life and compare that with populations with varying degrees of visual impairment as well as the general population.

## Data Availability

Data analysed during this study are included in this published article

## References

1. Lockley SW, Dressman MA, Licamele L, et al. Tasimelteon for non-24-hour sleep–wake disorder in totally blind people (SET and RESET): two multicentre, randomised, double-masked, placebo-controlled phase 3 trials. Lancet. 2015;386(10005):1754–1764. doi:10.1016/s0140-6736(15)60031-9.

2. Kobau R, Safran MA, Zack MM, Moriarty DG, Chapman D. Population tracking of perceived physical and mental health over time. Health Qual of Life Outcomes. 2004;2(1):1–37. doi:10.1186/1477-7525-2-40.

3. Centers for Disease Control and Prevention. Quality of life as a new public health measure--Behavioral Risk Factor Surveillance System, 1993. MMWR Morb Mortal Wkly Rep. 1994;43(20):375–380.

4. Crews JE, Chou C-F, Zhang X, Zack MM, Saaddine JB. Health-Related Quality of Life Among People Aged ≥65 Years with Self-reported Visual Impairment: Findings from the 2006–2010 Behavioral Risk Factor Surveillance System. Ophthalmic Epidemiol. 2014;21(5):287–296. doi:10.3109/09286586.2014.926556.

5. Argyriou K, Kapsoritakis A, Oikonomou K, Manolakis A, Tsakiridou E, Potamianos S. Disability in Patients with Inflammatory Bowel Disease: Correlations with Quality of Life and Patient’s Characteristics. Can J Gastroenterol Hepatol. 2017;2017:1–11. doi:10.1155/2017/613810.

6. Zheng K, Zhang S, Wang C, Zhao W, Shen H. Health-Related Quality of Life in Chinese Patients with Mild and Moderately Active Ulcerative Colitis. Plos One. 2015;10(4). doi:10.1371/journal.pone.0124211.

7. Lee PP, Spritzer K, Hays RD. The Impact of Blurred Vision on Functioning and Well-being. Ophthalmology. 1997;104(3):390–396. doi:10.1016/s0161-6420(97)30303-0.

8. Rajaratnam SM, Polymeropoulos MH, Fisher DM, et al. Melatonin agonist tasimelteon (VEC-162) for transient insomnia after sleep-time shift: two randomised controlled multicentre trials. Lancet. 2009;373(9662):482–491. doi:10.1016/s0140-6736(08)61812-7.

9. Groessl EJ, Liu L, Sklar M, Tally SR, Kaplan RM, Ganiats TG. Measuring the impact of cataract surgery on generic and vision-specific quality of life. Qual Life Res. 2012;22(6):1405–1414. doi:10.1007/s11136-012-0270-z.

10. Polack S, Eusebio C, Mathenge W, et al. The Impact of Cataract Surgery on Health Related Quality of Life in Kenya, the Philippines, and Bangladesh. Ophthalmic Epidemiol. 2010;17(6):387–399. doi:10.3109/09286586.2010.528136.

11. Cahill M, Banks A, Stinnett S, Toth C. Vision-related quality of life in patients with bilateral severe age-related macular degeneration. Ophthalmology. 2005;112(1):152–158. doi:10.1016/j.ophtha.2004.06.036.

12. Tsai S-Y, Chi L-Y, Cheng C-Y, Hsu W-M, Liu J-H, Chou P. The Impact of Visual Impairment and Use of Eye Services on Health-Related Quality of Life among the Elderly in Taiwan: The Shihpai Eye Study. Qual Life Res. 2004;13(8):1415–1424. doi:10.1023/b:qure.0000040791.87602.fe.

13. Chia E-M, Wang JJ, Rochtchina E, Smith W, Cumming RR, Mitchell P. Impact of Bilateral Visual Impairment on Health-Related Quality of Life: the Blue Mountains Eye Study. Invest Ophthalmol Vis Sci. 2004;45(1):71. doi:10.1167/iovs.03-0661.

14. Stelmack J. Quality of Life of Low-Vision Patients and Outcomes of Low-Vision Rehabilitation. Optom Vis Sci. 2001;78(5):335–342. doi:10.1097/00006324-200105000-00017.

15. Parrish R. Visual impairment, visual functioning, and quality of life assessments in patients with glaucoma. Am J Ophthalmol. 1997;123(4):578. doi:10.1016/s0002-9394(14)70205-3.

